# Cytokine response following perturbation of the cervicovaginal milieu during HPV genital infection

**DOI:** 10.1101/2021.02.10.21251486

**Authors:** Christian Selinger, Massilva Rahmoun, Carmen Lia Murall, Claire Bernat, Vanina Boué, Marine Bonneau, Christelle Graf, Sophie Grasset, Soraya Groc, Jacques Reynes, Christophe Hirtz, Nathalie Jacobs, Samuel Alizon

## Abstract

Human papillomaviruses (HPVs) are the most oncogenic viruses known to human, causing nearly all cervical cancers worldwide. Highly prevalent in young, sexually active women, most HPV infections are cleared within 3 years, and only a minority of those infections persist and lead to cancer later in life. To better characterize the immuno-modulatory impact of early HPV infection and more generally perturbations of the cervicovaginal milieu, we measured changes in a panel of 20 cytokines, known as highly dynamic effector molecules implicated in cell signaling. We analyzed 92 cervicovaginal samples collected from young, sexually active women who were tested for or diagnosed with HPV, chlamydia, and bacterial vaginosis. Also, symptoms associated with genital inflammation and infection were collected through self-reporting.

Following a parsimonious multi-factor modeling approach, our statistical analyses revealed that increased IL-1Alpha and IL-12/IL-23p40 concentrations were associated with HPV infection. Cytokine network analysis further highlighted the role of IL-1Alpha and macrophage inflammatory proteins (MIP-3Alpha) in HPV-associated immuno-modulation.

## Introduction

Human papillomaviruses (HPVs) are the most oncogenic viruses known to human, causing nearly all cervical cancers worldwide, as well as a significant fraction of other anogenital and oropharyngeal cancers.^1^ Even though a majority of sexually active women acquire a genital HPV infection, most cervical HPV infections will be cleared by the immune system within 3 years^2,3^ or lead to asymptomatic infections. The major risk factor for the development of high-grade cervical lesions and cervical cancer is the persistence of high-risk (HR) HPV type infection.^4^ Numerous studies on the HPV life cycle and pathogenesis show a broad spectrum of immune evasion strategies the virus uses to survive in its host.^5^

HPVs target the keratinocytes in the female genital tract and follow the differentiation program of these cells from the para-basal cells until the upper epithelial layers where virions are released. In these layers, immune cells have limited access, thereby facilitating virus immune escape. The cervicovaginal mucosa is known to perform a pivotal role in the defense against infectious and inflammatory processes thanks to an interplay between hormones, resident microbiota, epithelial, and immune cells. In particular, cervical keratinocytes express several toll-like receptors (TLRs) that recognize pathogen-associated molecular patterns and initiate innate and adaptive immune responses.^6^ However, given their slow replication cycle, HPVs are capable of maintaining low expression levels of viral proteins. They can also alter host gene expression (such as chemokines, adhesion molecules, and TLRs) or impair protein function and trafficking by dysregulating keratinocyte interferon (IFN) response and other antiviral pathways.^7–9^ Additionally, interference with antigen processing in HPV infections has been described and can contribute to impeding detection of infected keratinocytes by cytotoxic T cells.^10^

Most HPVs studies focus on chronic infections and cancer and there is a pronounced lack of data for non-persistent (also called “incident”) infections.^11^ The goal of this study is to analyze the cytokine-mediated response to HPV genital infections in healthy young women and describe the interplay, if any, between local cytokine secretion, HPV infection, and, more generally, infection-related perturbations to the cervicovaginal milieu.

The data originates from the PAPCLEAR clinical study^12^, which follows a population of sexually active, 18 to 25 year old women. We study a large panel of 20 soluble cytokines relevant to anti-viral responses and that fall into 4 groups: interleukins (IL-1Alpha, IL-5, IL-8, IL-10, IL-12p70, IL-12/IL-23p40, IL-15, IL-17A, IL-17A/F, IL-17E/IL-25, IL-18), interferons (IFN-Alpha2a, IFN-Beta, IFN-Gamma), TNF-Alpha (as a member of the TNF family), and chemokines (IP-10, MCP-1, MIP-1Alpha, MIP-3Alpha, and MIP-3Beta). These were selected for their role in innate or adaptive immunity, and because of their pro-inflammatory or anti-inflammatory activity. Here, we describe their cervicovaginal secretion profile.

Using statistical single-factor tests we found that increase in IL1-Alpha and IL-12/IL-23p40 levels is associated with HPV infection, whereas increased MIP-1Alpha is associated with HR HPV genotype infections only. To take into account additional factors pertaining to perturbations of the cervicovaginal milieu, we developed a model selection approach. Results indicate a broad cytokine response to chlamydia (IP-10, MIP-3Alpha, MCP-1, TNF-Alpha, MIP-3Beta, IL-15), and bacterial vaginosis (BV) (IL-17A/F, IL-8, IL-1Alpha, IL-15). However, cytokine response associated more specifically with changes in HPV infection are more limited (IL-1Alpha, IL-12/IL-23p40, IP-10, MIP-3Alpha). Our results from multi-factor analysis indicate that infections with HR HPV genotypes are associated with increased IP-10, IL-5 and MIP-1Alpha. Overall, our multivariate modeling approach provides a detailed description of cytokine expression profiles with great explanatory power in young women’s genital tract in the context of HPV and other genital infections.

## Results

### Study participants

Clinical and demographic information of the cohort has been described previously.^13^ For the subset of participants in this analysis the median age was 21 years (interquartile range: 21-23 years), and 5 percent of the study participants presented atypical squamous cells of undetermined significance (ASC-US) and 4% were diagnosed low-grade squamous intraepithelial lesions (LSIL). Forty out of 92 participants were HPV negative. The predominant HPV subtype was HPV-53 (13%), followed by HPV-51 (12%) and HPV-66 (10%). Two out of 92 participants had BV, both co-infected with HPV, and among the three participants with chlamydia diagnosis, two were co-infected with HPV.

### Genital cytokines and prevalent HPV infections

The lower limits of detection for the cytokine concentrations ranged between 10^-5.8 and 10^0.8 pg/ mL (see supplementary table 1). The distribution of normalized cytokine concentrations varies within four orders of magnitude in terms of median values and only a limited number of cytokines (MIP1-Alpha, IL-10, and IFN-Beta) are undetectable in 6-7% of the samples (Figure S1). Statistical tests suggest that several cytokines are not normally distributed (cytokines with low Shapiro-Wilk p-values, Figure S2) therefore we conducted suitable statistical single-factor tests relevant for HPV infections (Figure S3). Our results reveal that an increase in IL1-Alpha and IL-12/IL-23p40 concentration is associated with HPV positivity and clearance, whereas increases in MIP1-Alpha, IL1-Alpha, and IL-12/IL-23p40 concentrations are significantly associated with infection with HR HPV genotypes compared to low-risk genotypes or no HPV infection (Figure 1).

**Table 1:** Covariates related to infections

| Covariate name | Data type | Levels | Description | Source |
| --- | --- | --- | --- | --- |
| HPV+ | binary | yes/no | DEIA positive for at least one HPV genotype | LT |
| Multiple HPV+ | binary | yes/no | DEIA positive for more than one HPV genotype | LT |
| Never HPV+ | binary | yes/no | Never any occurrence of HPV infection in the past | QIV |
| HPV Pattern | categorical | 0_0, 0_1,<br>1_0, 1_1 | A_B means infection status A at visit 1 and B at visit 2; 0=DEIA negative for t at least one genotype; 1=DEIA positive for at least one genotype | LT |
| High risk HPV+ | binary | yes/no | yes: HR HPV genotype 16, 18, 31, 33, 35, 39, 45, 51, 52, 56, 58, 59<br>no: HPV negative or non-high-risk HPV positive | LT |
| Vaginosis+ | binary | yes/no | Any bacterial vaginosis diagnosis during past three months | QIV |
| Chlamydia+ | binary | yes/no | Either tested in past three months, if not, then tested at inclusion visit | STI screening at inclusion visit |
| Urinary tract infection+ | binary | yes/no | Any urinary infection diagnosis during past three months | QIV |
| Fungal Infection+ | binary | yes/no | Any fungal infection diagnosis during past three months | QIV |
| Genital infection+ | binary | yes/no | told by a health professional during your lifetime (and last three months) | QIV |
| Itching+ | binary | Yes/no | Any itching during past two weeks months | QIV |
| Vaginal odors+ | binary | yes/no | Any unusual vaginal odors during past two weeks | QIV |
| Douching+ | binary | yes/no | Any douching during past two weeks | QIV |
| Pelvic inflammation+ | binary | yes/no | Any pelvic inflammation diagnosis during past three months | QIV |
| Vaginal secretions+ | binary | yes/no | Any vaginal secretion during past two weeks | QIV |

Abbreviations: LT=laboratory testing, QIV= questionnaire at inclusion visit

**Figure 1:**
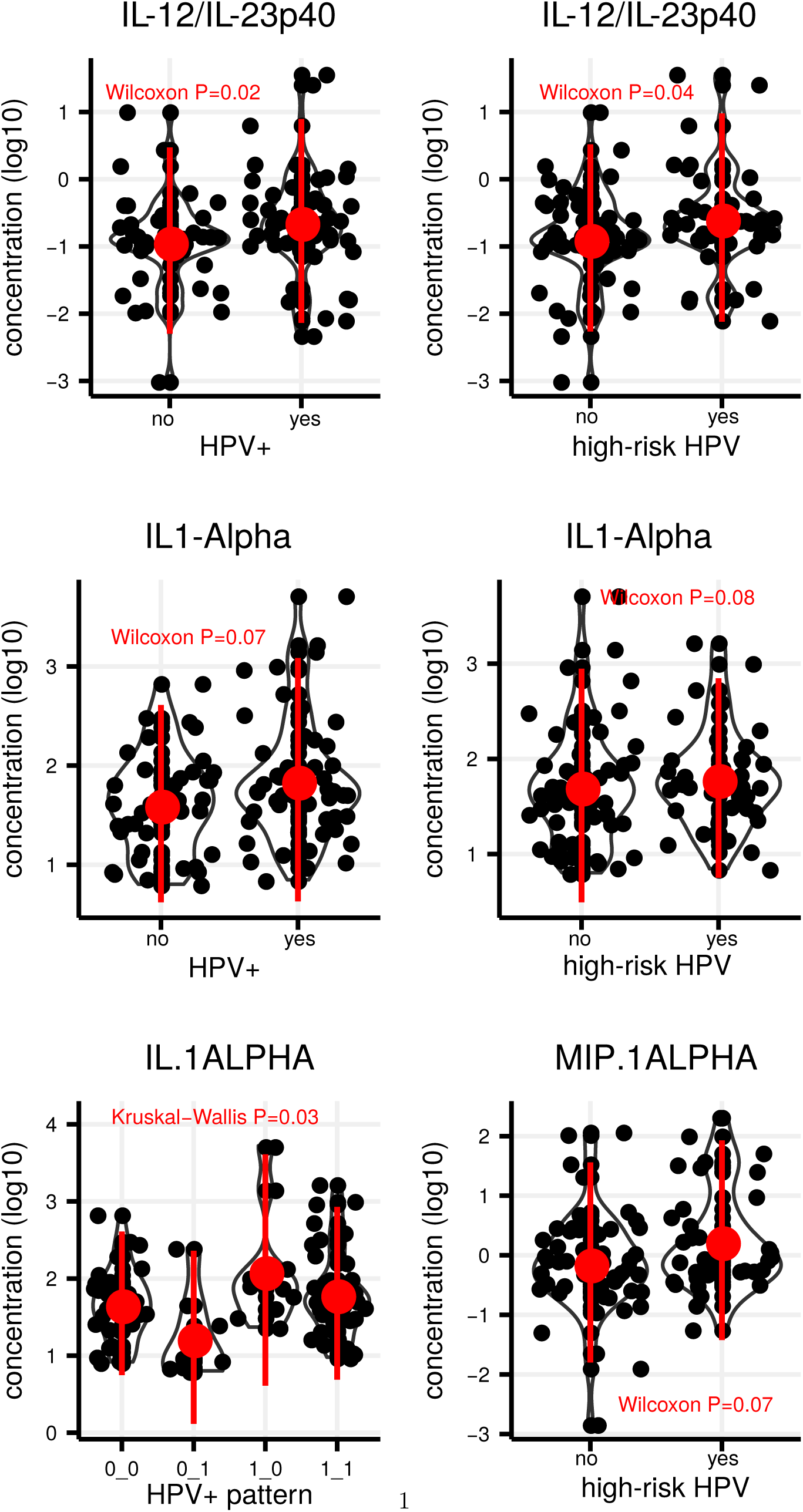
Single-factor statistical tests for cytokine changes with respect to HPV infection status. Black dots show data, the red dot is the median, and the red line the standard deviation for each group. Tests were chosen according to normality (see Figure S2). We show only results with p-value less or equal 0.1.

### Interplay between infection-related factors and cytokine response

To assess whether infection-related covariates produce a more generic cytokine response, we exhaustively tested 15 factors using linear models (see Material and Methods). Filtering models with at least one statistically significant covariate suggests that models combining between 10 and 12 factors were more likely to reveal significant associations (Figure S4).

The model selection approach used to determine the most parsimonious combination of covariates (Figure 2A) shows a broad panel of cytokines increasing in response to chlamydia (IP-10, MIP-3Beta, MCP-1, TNF-Alpha, MIP-3Alpha, IL-15) and, to a lesser extent, to BV (IL-17A/F, IL-8, IL-1Alpha, IL-15). Regarding, more specifically, the presence of an HPV infection, we find an increase of IL-1Alpha and IL-12/IL-23p40, as well as a reduction of IP-10 and MIP-3Alpha.

**Figure 2:**
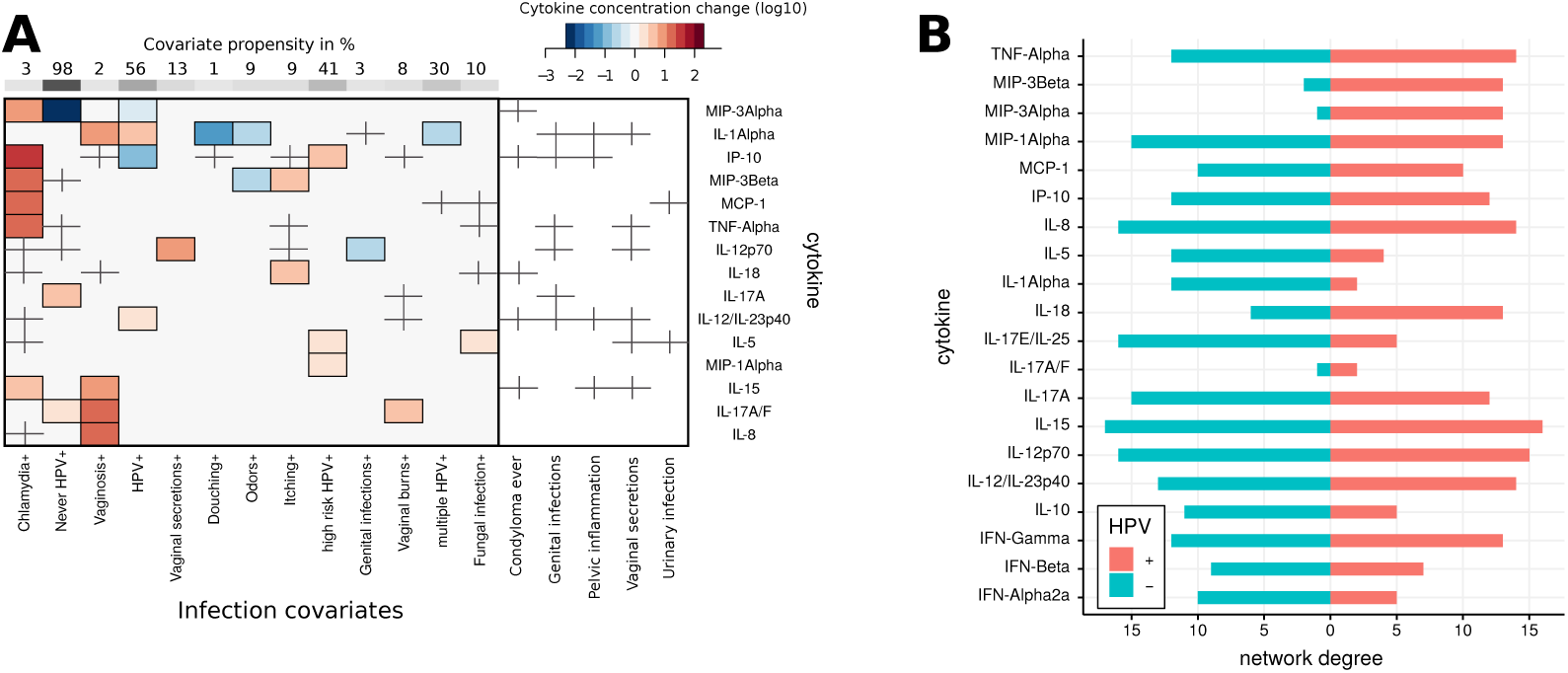
Panel A: The heatmap shows for each cytokine the most parsimonious linear model with increasing (red) and decreasing (blue) cytokine levels depending on infection covariate levels (columns, negative or absence considered as reference level). Crosses indicate the presence of a covariate in the selected model without being itself statistically significant. The propensity score indicates the percentage of a covariate level within the study population. Panel B: Degree distribution of cytokine correlation graphs for HPV positive (red) and HPV negative (turquoise) samples. Correlations were calculated using Pearson correlation p-values, the degree for a particular cytokine is defined as the number of correlated (*P <* 0.05) cytokines.

Infection with HR HPV genotypes is associated with increased levels of IP-10, IL-5, and MIP-1Alpha. Also, a majority of parsimonious models comprise covariates related to inflammatory perturbations (e.g. genital and urinary infections, pelvic inflammation). Although they are not significant by themselves, their contribution to the model selection process (Figure 2A, crossed fields) helps disentangle whether effects are related to HPV infection or not. E.g. the increase in IL-1Alpha for HPV positive participants was also impacted, but not significantly, by pelvic inflammation and genital infection status, and vaginal secretions.

### Cytokine network motifs

To investigate the relationship between cytokines, we first estimated correlation networks based on HPV infection status. The network degree of a particular node, which corresponds to a cytokine, is given by the number of other cytokines it is significantly correlated with. The most striking difference between the HPV-positive and the HPV-negative network was found for MIP-3Beta and MIP-3Alpha (Figure 2B). For HPV positive participants, these two cytokines were highly correlated with most other cytokines in the panel, whereas for HPV negative participants, significant correlations were largely absent. On the other hand, the number of cytokines correlated with IL-5 and IL-1Alpha increased for the HPV negative group when compared to HPV positive subjects (Figure 2B).

To evaluate the relationship between cytokines beyond correlations, we tested for changes in pairwise difference between cytokines using covariate combinations that have been selected in the multi-variate analysis from the preceding paragraph. Unlike correlations, which concern parallel trends, this approach allows to test whether the difference in concentration between two cytokines changes according to infection conditions. We found that MIP-3Alpha and IL-1Alpha (Figure 3) have highly contrasting roles in the cytokine network when HPV infection status changes. In particular, an IL-1Alpha increase is strongly associated with a MIP-3Alpha decrease, but with decreasing difference between the two cytokines (Figure 3, HPV+ panel with inward pointing arrows). On the other hand, the concurrent increase of IL-12/IL-23p40 and IL1-Alpha occurred with significantly increasing difference (Figure 3, HPV+ panel with outward pointing arrows). Similarly, the (self-reported) absence of HPV infection history prior to entry in the clinical study shows significant differences between IL-17A/F, IL-12p70 and IL-17E/IL-25. Interestingly, other covariates involving HPV status (multiple infections, HR-HPV genotypes) do not display significant network motifs, although each of them has at least one significantly different cytokine when tested alone. Chlamydia and BV covariates show the most striking network motifs, albeit with low propensity in the study population (3% and 2%, respectively).

**Figure 3:**
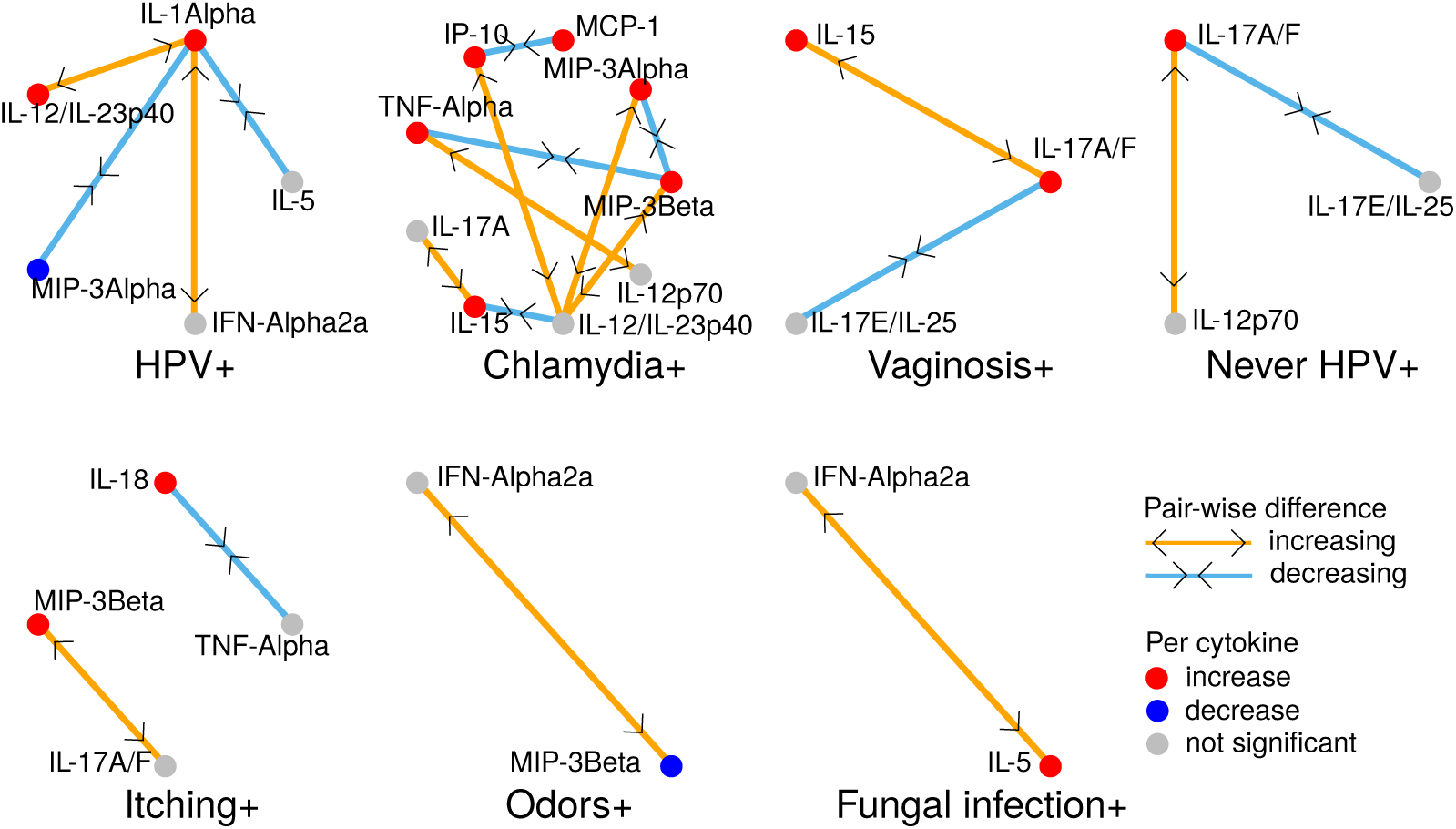
Network motifs of significant differences between cytokine concentrations. For each covariate, nodes in the motif correspond to cytokines that were included in at least one univariate model containing the covariate (Figure 2A) or had at least one significant difference with such a cytokine. Cytokines that significantly increased or decreased (resp. were not significant) in the per cytokine analysis (Figure 2A) are in red or blue (resp. grey). Increasing or decreasing differences between cytokines are marked by orange (with outward pointing arrows) or light blue edges (with inward pointing arrows).

## Discussion

Incident genital HPV infections in young women, especially in the absence of abnormal cervical cytology, have rarely been studied *in vivo*.^14,15^ To further our understanding of the natural history of such infections, it is necessary to characterize the host immune response, as persistent HPV infections have been hypothesized to evade the host immune system by modulating cytokine production and impairing pro-inflammatory responses.^7,16^

Determining signatures of cervicovaginal cytokine response to HPV infection has not yet reached consensus in the research community.^17–20^ The challenges are numerous and at various scales, ranging from cellular heterogeneity of solid tissue and non-detectability, to demographic and behavioral determinants. The PAPCLEAR cohort^12^ had been set up prospectively to yield a fairly homogeneous study population in terms of age, sexual activity and thus exposure history prior to enrollment. Incidentally, it also turned out to be well-balanced in terms of HPV infections such that it constitutes an opportunity to measure local immune mediators, such as cytokines, taking also into account covariates pertaining to microbial perturbations of the cervicovaginal environment. These include HPV infections, BV, chlamydia but also associated self-reported symptoms (odors, itching, etc.).

A high concentration of IL-1Alpha pro-inflammatory cytokine was detected in the cervicovaginal environment of our healthy cohort and it was significantly associated with HPV infection and clearance. This is consistent with the fact that this cytokine is produced by keratinocytes and is a crucial mediator of the antigen recognition process by Langerhans cells that migrate then to draining lymph nodes. This result is in line with reports^21,22^ who detected significantly higher levels of inflammatory mediators including IL-1Alpha in vaginal swabs collected from sexually active compared to sexually inactive girls.

Although we can statistically corroborate the association of several cytokines (IL-1Alpha, IL-12/IL-23p40, MIP-1Alpha) with HPV infection,^18^ these were not confirmed by a recent study^19^ focusing on the link between persistence of HPV infection and pro-inflammatory cytokines (including IL-1Alpha, IL-10, IL-17, and TNF-Alpha). This difference could be due to bias in the ethnic origin of the participants, sampling method (self-collection in Softcups versus gynecologist Weck-Cel sponge collection in our study) or concentration normalization methods. Note also that cervical IL-1Alpha concentrations vary during the menstrual cycle^23^ with a potential protective role against bacterial invasion into the uterine cervix.^24^

We also detected novel significant association between the presence of HR-HPV genotypes and the MIP-1Alpha chemokine, as well as the IL-12/IL-23p40 interleukin. IL-12 is an important regulator of T-cell functions, known to promote T-helper-1 cell development, and IL23 is a major inducer of IL-17. These data are consistent with the reported locally immunosuppressive function of IL-17 in HPV-associated premalignant disease.^25,26^

The assessment of cytokine responses revealed that specific soluble immune mediators, mainly chemokines (IP10, MIP-3Beta, MCP-1, and MIP-3Alpha) are positively associated with chlamydia infection. Chemokines are small cationic molecules that link innate and adaptive immune responses by attracting effector cells to the site of infection. Chemokines such as MIP-3Alpha are known to act as an antimicrobial agent.^27^ Similarly, the Th1-associated chemokines IP-10 and MCP-1 have been reported to be important in immunity against chlamydia.^28,29^ Our results point also to the known involvement of NK-cells via the action of IL-15 and TNF-Alpha^30^, which we found both associated with significant effects.

BV is the most common vaginal infection in women of reproductive age and is associated with an increased risk of acquiring sexually transmitted infections including HPV infections.^31^ Among the cytokines, we found that a BV diagnosis was significantly associated with the presence of IL-1Alpha, consistently with earlier results.^32^ Overall, our findings are consistent with the cytokine signatures defined by Masson and colleagues^33^ with chlamydia being associated with the highest genital cytokine levels and BV-infected women displaying a mixed genital cytokine profile, with increased proinflammatory cytokine concentration.

We extended prior analyses by investigating the interplay between cytokines depending on microbial perturbations in two ways. First, our statistical analysis was able to integrate combinations of covariates in order to yield for each cytokine the most parsimonious descriptors with an optimal trade-off between predictive power and degrees of freedom. Second, correlation network analysis confirmed the central role played by IL-1Alpha. Network motif analysis showed that IL-12/IL-23p40 not only increased concurrently with IL-1Alpha, but that their difference was also increasing, suggesting non-linear relationships.

Functional interpretation remains a major limitation of our study, since pathway analysis based on expression patterns^34^ typically requires several hundred transcripts from host mRNA, and has also to account for microbial community states.^35^ We deliberately limited our analysis to covariates related to microbial perturbations, since extending this approach to other covariates (e.g. vaccination or sexual behavior) would likely result in model overfitting, given the size of the cohort.

A perspective of this study will be to perform longitudinal analyses on the PAPCLEAR cohort data. It will be particularly informative to investigate how the dynamics of the cytokines strongly associated with HPV infection (e.g. IP-10, MIP-3Alpha, and IL-1Alpha) correlates with viral load dynamics. Additionally, the vaginal microbiome profiling of our cohort could shed light on the impact of chlamydia or BV in an HPV context. Taken together, our analysis revealed novel cytokines involved in local immune responses to acute HR-HPV infection and confirmed the central role played by IL-1Alpha more generally toward HPV infection.

## Materials and Methods

### Participant enrollment and sampling

The PAPCLEAR study is a mono-centric longitudinal study run at the Montpellier STI detection center (CeGIDD). Its main objective is to decipher the kinetics of HPV genital infections in young women. The enrollment criteria and study design have been described previously.^12^

During the inclusion visit, participants filled-out online health and lifestyle questionnaires that capture a range of demographic, clinical, reproductive, and behavioral variables. Cervicovaginal secretions were collected using Weck-Cel sponges (Beaver-Visitec International) placed directly into the cervical os for approximately 1 min. Sponges were then transferred into a Salivette® (Sarstedt) device and centrifugated at 1500rpm for 5 min at 4°C after the addition of 175µL of phosphate-buffered saline (PBS). Supernatants were separated into 50µL aliquots and stored at −80°C.

### Detection of HPV DNA and other infections

The protocols used for HPV detection and genotyping rely on the DEIA and LiPa25 tests as described previously.^13^ The presence of chlamydia infection was tested by PCR following the STI detection center standard procedures. Diagnosis for BV, and urinary tract, fungal, or genital infections were performed by the clinician or mid-wife during the gynecological visit.

Cytokine concentrations were analyzed at the first (inclusion) visit of 103 participants. For HPV testing and genotyping, we also used data from the second visit, a month later, to define more robust infection patterns and rule out transient HPV carriage, as detailed in an earlier study.^13^

### Measurement of cervicovaginal cytokine concentrations

The MSD U-plex Biomarker Group 1 (human) from Meso Scale Discovery (MSD, Rockville, Maryland, USA) was used to measure 20 analytes divided into two panels: IFN-Gamma, IL-5, IL-8, IL-10, IL-12p70, IL-17A, TNF-Alpha, IP-10, MCP-1, MIP-1Alpha, and IFN-Alpha2a on one plate; IL-1Alpha, IL-12/IL-23p40, IL-15, IL-18, MIP-3Alpha, MIP-3Beta, IL-17A/F, IL-17E/IL-25, and IFN-Beta on a second plate. For each panel, we used 25µL of specimen, according to the manufacturer’s instructions. Analyses were performed on a MESO QuickPlex SQ 120 reader.

### Measurement of total protein

To normalize cytokine quantitation, total protein content was measured in the same samples using the Invitrogen Qubit protein assay kit (Thermo Fisher Scientific) following the manufacturer’s instructions.

### Statistical analysis

To calculate cytokine sample concentrations from the standard curves, we used a four-parameter logistic regression model^36,37^ implemented in R with the package *minpack*.*lm*. In order to avoid non-detectability as a confounding factor, we only kept samples for which all 20 cytokines were detectable across all participants, resulting in 92 (of initially 103) samples. We divided each concentration by the total amount of protein measured in each sample, before transforming the normalized cytokine concentration to log10 scale. We tested each cytokine distribution for normality using the Shapiro-Wilk test. We tested for differences in cytokine concentration associated with changes regarding HPV infections using parametric (t-test for normally distributed cytokines) and non-parametric tests (else, with Wilcoxon test for all binary covariates and Kruskal-Wallis test for covariates with more than two levels).

We then applied linear regression models with fixed effects for combinations of covariates based on the survey conducted during the inclusion visit resulting in a set of 15 factors pertaining to infections (see Table 1 for a complete list). In these models, we were adjusting for the standard curve calibration parameter setting and the total protein quantity used for normalization. For each cytokine, we tested all 2^15^ covariate combinations, and among the models with at least one statistically significant factor and normally distributed residuals (using a Shapiro-Wilk test criteria), we chose the one with the lowest Akaike Information Criterion (AIC) value to select the most parsimonious model.

Furthermore, associations between groups of cytokines relating to infection covariates were investigated by comparing correlation networks. Given the set of 20 nodes (i.e. cytokines), we would draw an edge between two nodes, whenever the correlation between respective concentrations was statistically significant (Pearson correlation with P<0.05). We then compared changes in degree distribution (i.e. the number of significant correlations of a cytokine with all other cytokines) between HPV positive and HPV negative samples.

In order to relate associations between cytokines to perturbations of the cervicovaginal milieu, we tested pairwise differences between cytokine concentrations with independent variables obtained from the model selection procedure of fixed-effect linear models above. This approach helped identify significant changes between cytokines depending on changes in infection covariates. To summarize our results, we visualized essential cytokine network motifs. For each covariate, nodes in the motif consist of cytokines that were included in at least one univariate model containing the covariate (Figure 2A) or had at least one significant difference with a cytokine doing so. Cytokines that significantly increased or decreased (resp. were not significant) in the per cytokine analysis (Figure 2A) are in red or blue (resp. grey). Increasing or decreasing differences between cytokines are marked by orange or light blue edges. To further highlight the increasing (resp. decreasing trend) of differences we added with outward (resp. inward) pointing arrows to the edges. The node position or the edge length within a motif are only part of the graphic layout and have no statistical meaning. Only motifs with at least one significant difference (i.e. edge) were plotted.

All analyses were performed in R (R Development Core Team (2019). R: A Language and Environment for Statistical Computing. Vienna, Austria: R Foundation for Statistical Computing) using packages lme4 (*glm*) and stats (*kruskal*.*test, t*.*test, shapiro*.*test, wilcox*.*test*). Scripts and data will be deposited in the Zenodo repository.

## Data Availability

Scripts and data will be deposited in the Zenodo repository.

## Ethics

The PAPCLEAR trial is promoted by the Centre Hospitalier Universitaire de Montpellier and has been approved by the Comité de Protection des Personnes (CPP) Sud Méditerranée I on 11 May2016 (CPP number 16 42, reference number ID RCB 2016-A00712-49); by the Comité Consultatif sur le Traitement de l’Information en matière de Recherche dans le domaine de la Santé on 12 July 2016 (reference number 16.504); and by the Commission Nationale Informatique et Libertés on 16 December 2016 (reference number MMS/ABD/AR1612278, decision number DR-2016–488). This trial was authorised by the Agence Nationale de Sécurité du Médicament et des Produits de Santé on 20 July 2016 (reference20160072000007). The ClinicalTrials.gov identifier is NCT02946346. All participants provided written informed consent.

## Acknowledgments

The authors acknowledge the IRD itrop HPC (South Green Platform) at IRD Montpellier for providing HPC resources that have contributed to the research results reported within this paper (URL: http://www.southgreen.fr).

We also acknowledge productive feedback on the manuscript by Nicolas Tessandier.

This work was supported by the European Research Council (ERC) under the European Union’s Horizon 2020 research and innovation program [grant agreement No 648963 to SA]. The sponsor had no role in study design, in the collection, analysis and interpretation of data, in the writing of the report and in the decision to submit the article for publication.

## Contributions

Conceived the study: SA, CLM, CH, NJ, JR

Revised the manuscript: SA, CS, MR, CLM, NJ

Performed the experiments: MR, CB, VB, Soraya G, SG, CG, MB

Interpreted the results: MR, CS, SA

Wrote the initial draft: CS, MR

Performed the statistical analysis: CS

## Supplementary Material

**Supplementary table 1:** Lower limits of cytokine concentration (log10)

|  |  |
| --- | --- |
| IL-10 | -4.75 |
| IL-15 | -3.48 |
| IL-5 | -2.63 |
| IL-12p70 | -3.29 |
| IL-17E/IL-25 | -4.19 |
| IL-17A/F | -2.56 |
| TNF-Alpha | -3.44 |
| IL-12/IL-23p40 | -3 |
| IFN-Alpha2a | -2.99 |
| IFN-Gamma | -4.27 |
| IFN-Beta | -5.8 |
| MIP-3Beta | -2.02 |
| IL-17A | -2.55 |
| MIP-1Alpha | -2.83 |
| MCP-1 | -0.82 |
| MIP-3Alpha | -2.31 |
| IL-1Alpha | 0.8 |
| IL-18 | -0.1 |
| IP-10 | -0.86 |
| IL-8 | 0.49 |

**Supplementary Figure 1:**
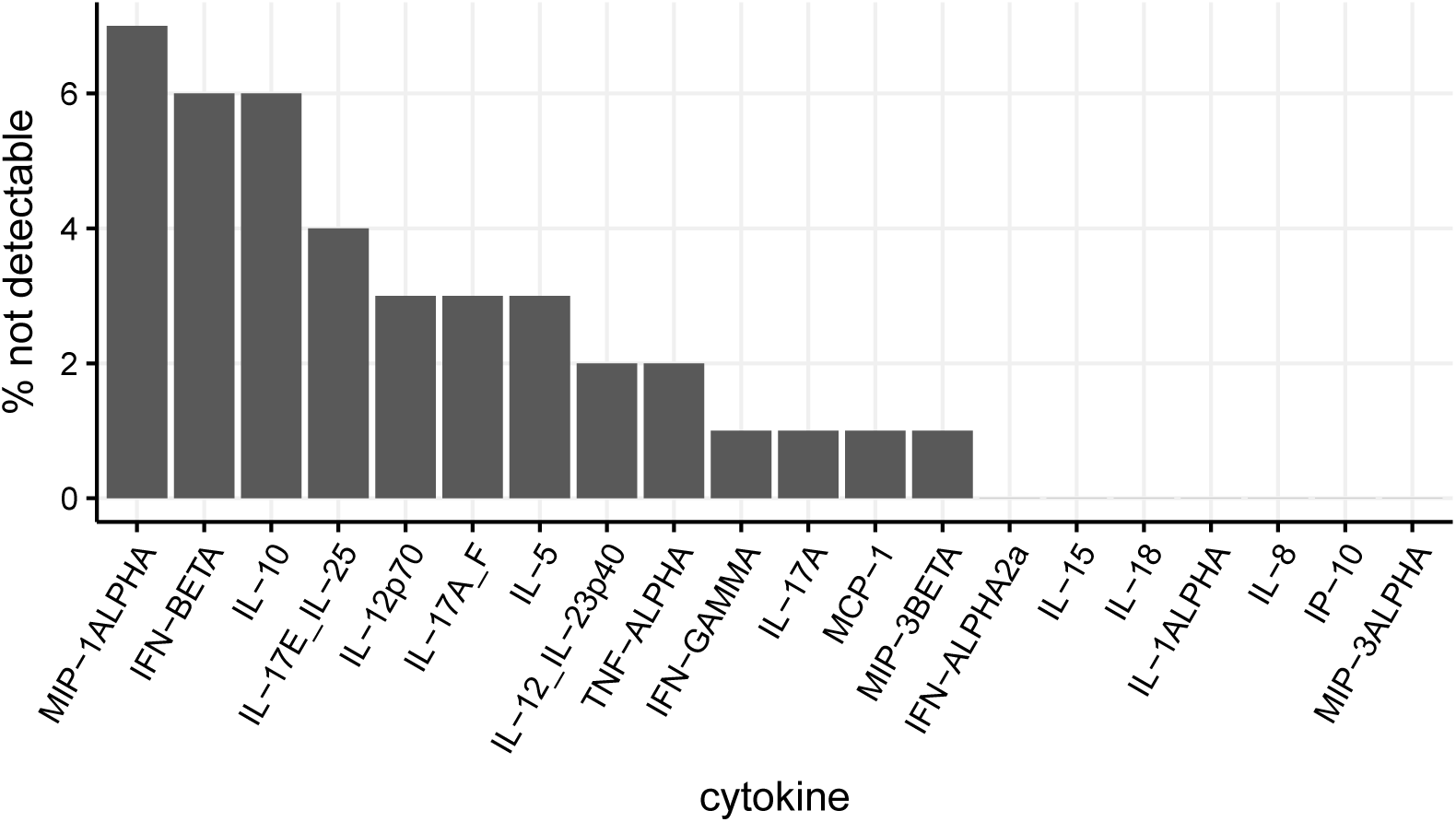
Detection propensity for cytokines in 103 initial samples.

**Supplementary Figure 2:**
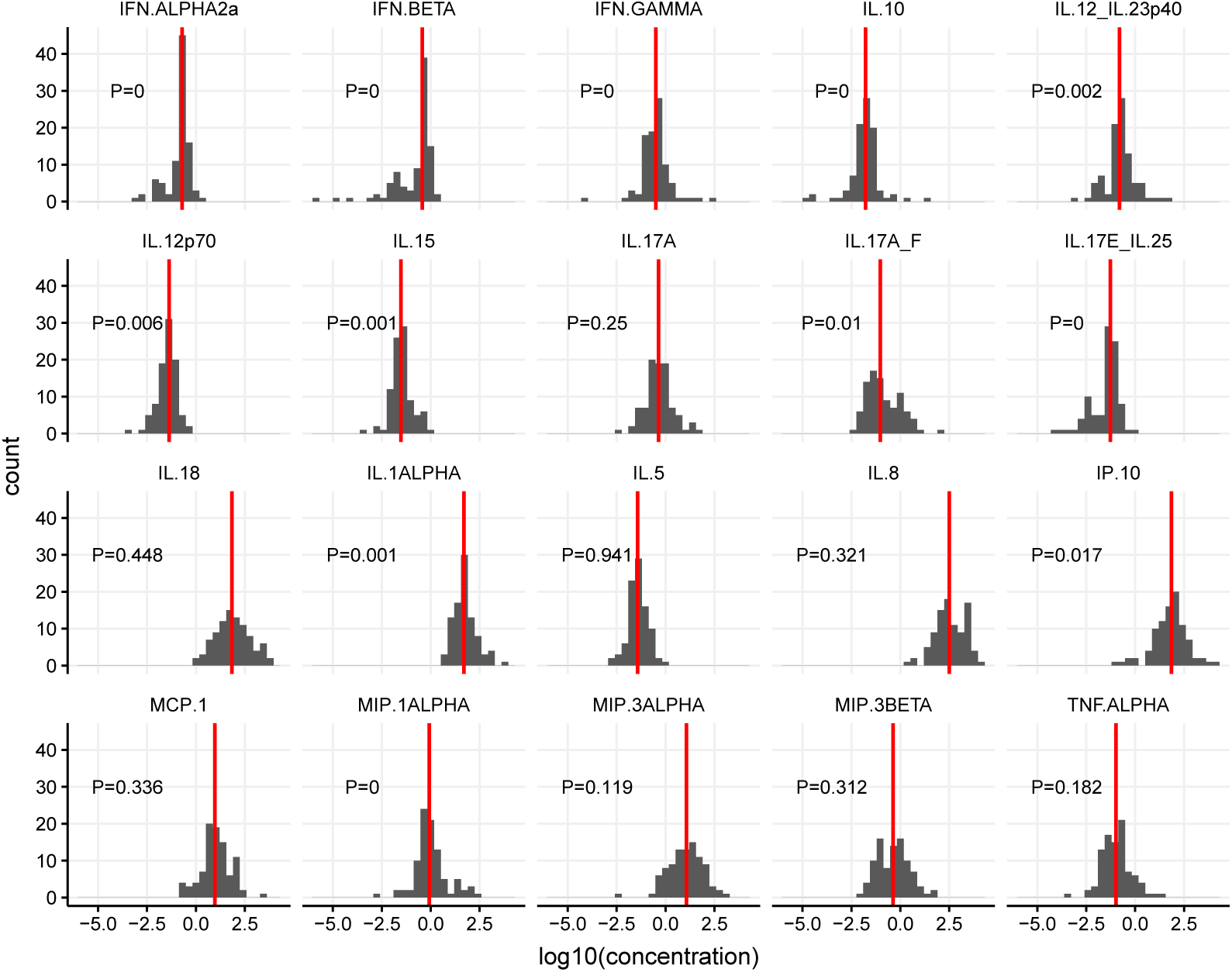
Distribution of normalized cytokine concentrations (log10) for complete observations with p-values from the Shapiro-Wilk test, P¡0.05 indicating non-Gaussian distribution.

**Supplementary Figure 3:**
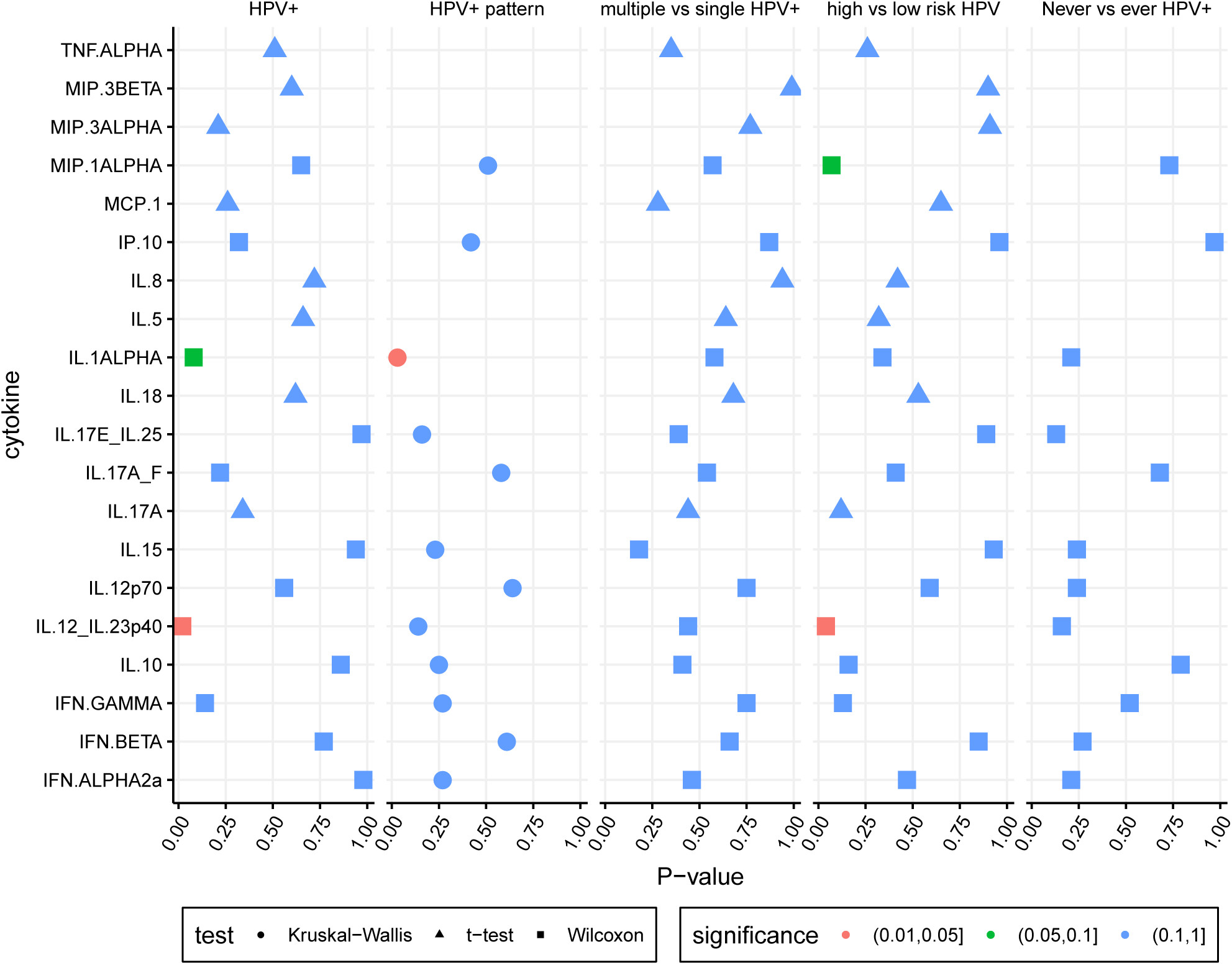
Results of statistical tests for HPV infection covariates using parametric (t-test for normally distributed cytokines) and non-parametric tests (Wilcoxon test for all binary covariates, Kruskal-Wallis test for covariates with more than two levels for non-Gaussian distributions).

**Supplementary Figure 4:**
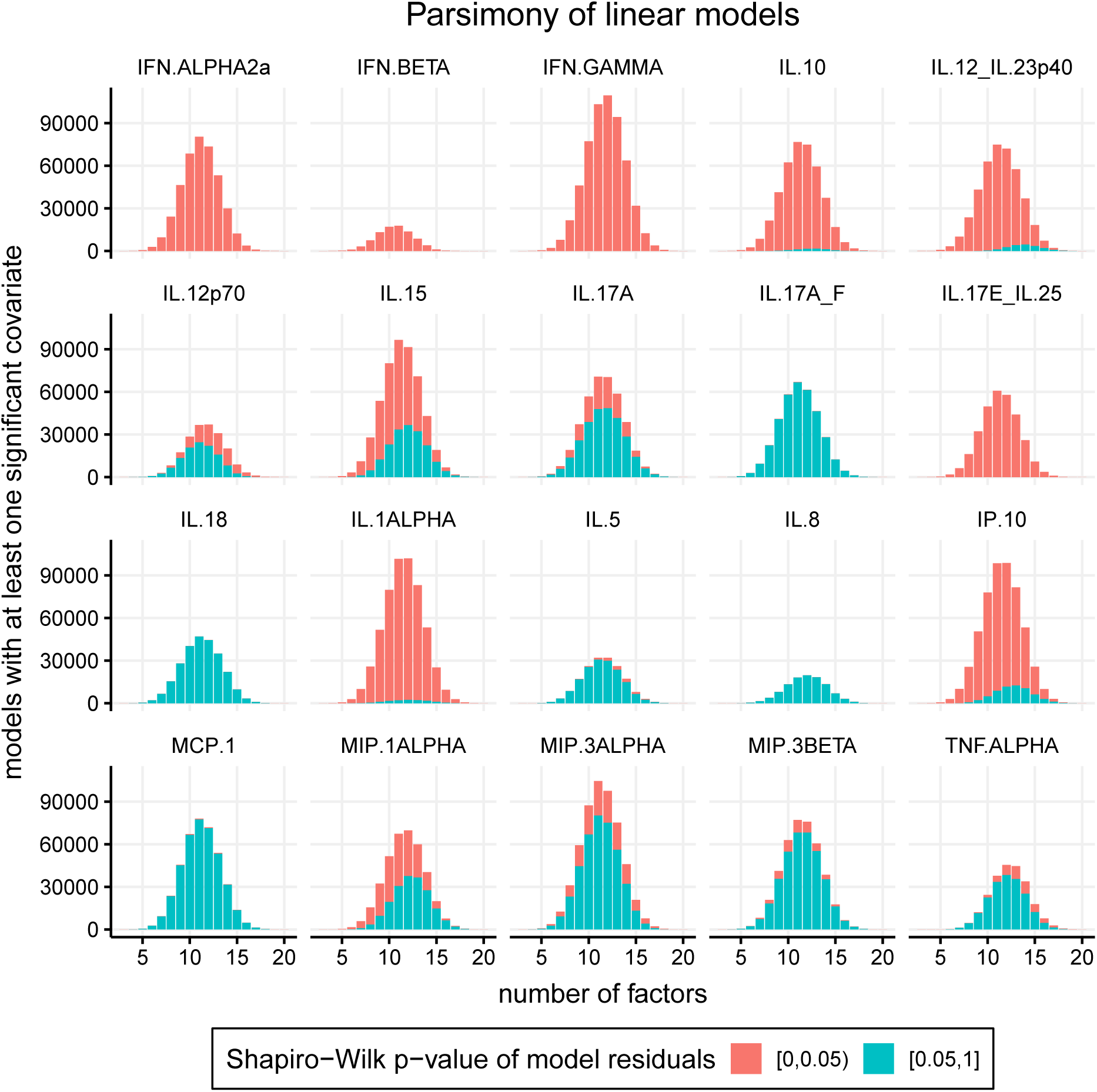
Number of linear models with at least one statistically significant covariate, with normally (turquoise) and not normally (red) distributed residuals depending on the number of covariates in each model.

## References

1. Martel, C. de, Plummer, M., Vignat, J. & Franceschi, S. Worldwide burden of cancer attributable to HPV by site, country and HPV type. International Journal of Cancer 141, 664–670 (2017).

2. Schiffman, M., Castle, P. E., Jeronimo, J., Rodriguez, A. C. & Wacholder, S. Human papillomavirus and cervical cancer. Lancet 370, 890–907 (2007).

3. Rodríguez, A. C. et al.. Rapid clearance of human papillomavirus and implications for clinical focus on persistent infections. J. Natl. Cancer Inst. 100, 513–517 (2008).

4. IARC working group on the evaluation of carcinogenic risks to humans: occupational exposures of hairdressers and barbers and personal use of hair colourants; some hair dyes, cosmetic colourants, industrial dyestuffs and aromatic amines. Proceedings. Lyon, France, 6-13 October 1992. IARC Monogr Eval Carcinog Risks Hum 57, 7–398 (1993).

5. Steinbach, A. & Riemer, A. B. Immune evasion mechanisms of human papillomavirus: An update. Int. J. Cancer 142, 224–229 (2018).

6. Nasu, K. & Narahara, H. Pattern Recognition via the Toll-Like Receptor System in the Human Female Genital Tract. Mediators of Inflammation 2010, (2010).

7. Amador-Molina, A., Hernández-Valencia, J. F., Lamoyi, E., Contreras-Paredes, A. & Lizano, M. Role of Innate Immunity against Human Papillomavirus (HPV) Infections and Effect of Adjuvants in Promoting Specific Immune Response. Viruses 5, 2624–2642 (2013).

8. Nunes, R. A. L., Morale, M. G., Silva, G.Á.F., Villa, L. L. & Termini, L. Innate immunity and HPV: friends or foes. Clinics (Sao Paulo) 73, e549s (2018).

9. Stanley, M. Immunology of HPV Infection. Curr Obstet Gynecol Rep 4, 195–200 (2015).

10. Leone, P. et al. MHC class I antigen processing and presenting machinery: organization, function, and defects in tumor cells. J. Natl. Cancer Inst. 105, 1172–1187 (2013).

11. Alizon, S., Murall, C. L. & Bravo, I. G. Why Human Papillomavirus Acute Infections Matter. Viruses 9, 293 (2017).

12. Murall, C. L. et al. Natural history, dynamics, and ecology of human papillomaviruses in genital infections of young women: protocol of the PAPCLEAR cohort study. BMJ Open 9, e025129 (2019).

13. Murall, C. L. et al. HPV cervical infections and serological status in vaccinated and unvaccinated women. Vaccine 38, 8167–8174 (2020).

14. Moscicki, A. B. et al. The natural history of human papillomavirus infection as measured by repeated DNA testing in adolescent and young women. J Pediatr 132, 277–284 (1998).

15. Zanotta, N. et al. Candidate Soluble Immune Mediators in Young Women with High-Risk Human Papillomavirus Infection: High Expression of Chemokines Promoting Angiogenesis and Cell Proliferation. PLoS One 11, e0151851 (2016).

16. Tummers, B. & Van Der Burg, S. H. High-Risk Human Papillomavirus Targets Crossroads in Immune Signaling. Viruses 7, 2485–2506 (2015).

17. Fernandes, A. P. M. et al. HPV16, HPV18, and HIV infection may influence cervical cytokine intralesional levels. Virology 334, 294–298 (2005).

18. Liebenberg, L. J. P. et al. HPV infection and the genital cytokine milieu in women at high risk of HIV acquisition. Nat Commun 10, 5227 (2019).

19. Shannon, B. et al. Association of HPV infection and clearance with cervicovaginal immunology and the vaginal microbiota. Mucosal Immunol 10, 1310–1319 (2017).

20. Moscicki, A.-B., Shi, B., Huang, H., Barnard, E. & Li, H. Cervical-Vaginal Microbiome and Associated Cytokine Profiles in a Prospective Study of HPV 16 Acquisition, Persistence, and Clearance. Front. Cell. Infect. Microbiol. 10, (2020).

21. Jespers, V. et al. Association of Sexual Debut in Adolescents With Microbiota and Inflammatory Markers. Obstet Gynecol 128, 22–31 (2016).

22. Ghosh, M. et al. Immune biomarkers and anti-HIV activity in the reproductive tract of sexually active and sexually inactive adolescent girls. Am J Reprod Immunol 79, e12846 (2018).

23. Boily-Larouche, G. et al. Characterization of the Genital Mucosa Immune Profile to Distinguish Phases of the Menstrual Cycle: Implications for HIV Susceptibility. J Infect Dis 219, 856–866 (2019).

24. Kanai, T. et al. Increased interleukin-1 and interleukin-1 receptor antagonist levels in cervical mucus in the ovulatory phase in comparison with the follicular phase. Gynecol Obstet Invest 43, 166–170 (1997).

25. Gosmann, C., Mattarollo, S. R., Bridge, J. A., Frazer, I. H. & Blumenthal, A. IL-17 suppresses immune effector functions in human papillomavirus-associated epithelial hyperplasia. J Immunol 193, 2248–2257 (2014).

26. Hede, D. V. et al. Human papillomavirus oncoproteins induce a reorganization of epithelial-associated γd T cells promoting tumor formation. PNAS 114, E9056–E9065 (2017).

27. Yang, D. et al. Many chemokines including CCL20/MIP-3α display antimicrobial activity. Journal of Leukocyte Biology 74, 448–455 (2003).

28. Belay, T. et al. Chemokine and Chemokine Receptor Dynamics during Genital Chlamydial Infection. Infection and Immunity 70, 844–850 (2002).

29. Poston, T. B. et al. Cervical Cytokines Associated With Chlamydia trachomatis Susceptibility and Protection. J Infect Dis 220, 330–339 (2019).

30. Radomski, N. et al. Chlamydia psittaci-Infected Dendritic Cells Communicate with NK Cells via Exosomes To Activate Antibacterial Immunity. Infect Immun 88, (2019).

31. Gillet, E. et al. Bacterial vaginosis is associated with uterine cervical human papillomavirus infection: a meta-analysis. BMC Infect Dis 11, 10 (2011).

32. De Seta, F., Campisciano, G., Zanotta, N., Ricci, G. & Comar, M. The Vaginal Community State Types Microbiome-Immune Network as Key Factor for Bacterial Vaginosis and Aerobic Vaginitis. Front Microbiol 10, 2451 (2019).

33. Masson, L. et al. Defining genital tract cytokine signatures of sexually transmitted infections and bacterial vaginosis in women at high risk of HIV infection: a cross-sectional study. Sex Transm Infect 90, 580–587 (2014).

34. Selinger, C. et al. Cytokine systems approach demonstrates differences in innate and pro-inflammatory host responses between genetically distinct MERS-CoV isolates. BMC Genomics 15, 1161 (2014).

35. Elovitz, M. A. et al. Cervicovaginal microbiota and local immune response modulate the risk of spontaneous preterm delivery. Nature Communications 10, 1305 (2019).

36. Dunn, J. & Wild, D. Chapter 3.6 - Calibration Curve Fitting. In The Immunoassay Handbook (Fourth Edition) (Wild, D.) 323–336 (Elsevier, Oxford, 2013).doi:10.1016/B978-0-08-097037-0.00022-1

37. O’Connell, M. A., Belanger, B. A. & Haaland, P. D. Calibration and assay development using the four-parameter logistic model. Chemometrics and Intelligent Laboratory Systems 20, 97–114 (1993).

